# Analyzing open-ended questions in research: A commonly used category selection methodology

**DOI:** 10.1101/2022.05.27.22275646

**Authors:** Luis D. Agosto Arroyo, Angerica Fitzmaurice, Zlatan Feric, David Kaeli, John Meeker, Carmen Velez Vega, Akram Alshawabkeh, José F. Cordero, Nancy R. Cardona-Cordero

**Affiliations:** School of Public Health, Medical Sciences Campus, University of Puerto Rico, San Juan, PR; Department of Gynecology and Obstetrics, University of Rochester, Rochester, NY; Department of Engineering, Northeastern University, Boston, MA; Department of Epidemiology and Biostatistics, College of Public Health, University of Georgia, Athens, GA; School of Public Health, University of Michigan, Ann Arbor, MI; Department of Computer Science, Northeastern University, Boston, MA

**Author notes:** corresponding author Nancy R. Cardona-Cordero. Contributions: Luis D. Agosto Arroyo - Conceptualization, Data curation, Formal analysis, Investigation, Methodology, Supervision, Writing – original draft, Writing – review & editing Angerica Fitzmaurice - Data curation, Investigation, Writing – review & editing Zlatan Feric - Data curation, Investigation, Methodology David Kaeli - Funding acquisition, Investigation, Supervision John Meeker - Funding acquisition, Investigation, Project administration Carmen Velez Vega - Funding acquisition, Investigation, Writing – review & editing Akram Alshawabkeh - Conceptualization, Funding acquisition, Investigation, Project administration, Supervision, Writing – review & editing José F. Cordero - Conceptualization, Funding acquisition, Investigation, Project administration, Supervision, Writing – review & editing Nancy R. Cardona-Cordero - Conceptualization, Funding acquisition, Investigation, Methodology, Project administration, Resources, Supervision, Writing – review & editing.

## Abstract

A closer examination of consumer product brands and how they are associated with levels of potential endocrine disrupting chemicals should be explored. The large number of brands available and changes in consumer preferences for certain brands makes it difficult to develop questionnaires that include all brands. Open-ended brand reporting questions are an option, but they bring challenges in identifying each brand given the multiple possibilities of variations in brand name reporting. We report a method for transforming product brand data reported as text to brand codes that allows quantitative analysis of brand use and its association with endocrine disrupting chemicals. We selected 14 consumer products to be included in our analyses. To evaluate commonly used brand selection, we used Cohen’s power calculations for two-sample t-tests in R (version 1.3.0). Considering a moderate effect size (Cohen’s d) of 0.5, each test will include the most used brand and the least used brand among the commonly used brands per product and visit. We compared how the commonly used brand selection differ per product in terms of the number of brands it selected, the total sample size and the power calculated by creating a correlation matrix and analyzing the relationship between power, commonly used brands, and brand usage. The correlation coefficient between the commonly used brand frequency of each visit approximated 0.99. From all products, fabric softener, conditioner, and lotion where the products that attained the highest power. The differences in brand use distributions per product provided an optimal environment for evaluating the performance of the commonly used brand selection methodology. It provides enough flexibility when selecting exposure groups that it could be applied to any open-ended questions, and it proves significantly useful when accounting for repeated measures.

## Introduction

Numerous brands are being developed continuously for the diverse personal consumer products marketed globally. These are products often used daily, weekly, or monthly, and use of different brands may vary subject to the product availability, a person’s socio demographic characteristics. Use of personal care or home cleaning products may contain fragrances and other additives that may expose people to harmful chemicals such as phthalates and other persistent organic pollutants (POPs). Studies have shown that consumer products can act as predictors of exposure for phenols, parabens, phthalates, and many other chemicals (1-3). These substances are also endocrine disrupting chemicals (EDCs) and have been associated with adverse reproductive health outcomes (4-6). A recent study (7) found that elevated urine levels of triclocarban and triclosan metabolites among participants who reported use of specific bar soap brands. While several brands listed triclocarban/triclosan as an additive in their bar soap, the levels were not listed and pointed out the importance of examining specific brands rather than considering all brands of bar soap together. This finding has led to a closer examination of consumer product brands and how they are associated with levels of potential EDCs in the environment. The multiple brands of commercial products present a major challenge to surveys because brand names vary locally, regionally, and globally. Developing surveys with a systematic list of personal care brands is complex and often not practical. We have used open ended questions in our product use survey and extracted the brand name. We are unaware that this approach has been used before in studies to assess human exposures and their potential health effects.

A challenge to this type of analysis is that the large number of brands available and changes in consumer preferences for certain brands makes it difficult to develop questionnaires that include all brands. Moreover, long lists of brands complicate the administration of questionnaires. Open-ended brand reporting is an option, but it brings challenges in how to identify each brand given the multiple possibilities of variations in brand name reporting. Moreover, for some personal care products there are many brands are available with varying degrees of market share which requires an examination of brands mostly used by consumers responding to the survey. Open-ended responses pose a major challenge when studies are aiming to target specific exposures in health sciences. For decades, closed questions have dominated psychological, social and health sciences for obvious reasons including a generally high-test score reliability as well as the possibility to standardize data collection and study comparisons (8). However, open ended prompts may be reliably coded and provide a more in-depth understanding of the exposure phenomena. Open questions may instead be used as predictors in statistical analyses rather than endpoint variables, providing and individual or collective variability in an exposure assessment of a possible salient issue (8, 9).

We report a method for transforming product brand data reported as text into brand codes that allows quantitative analysis of brand use and its association with EDCs levels and its possible association with a health condition. We also describe an approach to develop categories of most used brands, that account to majority of exposures, when there are many brands involved.

## Methods

### Data source

We used data obtained by the Puerto Rico Testsite for Exploring Contaminations Threats (PROTECT) study. PROTECT is a Superfund Research Center that examines the effects of POPs on adverse pregnancy outcomes through a prospective maternal-child cohort that evaluates fate and transport of environmental contamination and its relationship with adverse pregnancy outcomes, such as preterm birth, in the northern karst area in Puerto Rico. Details of this cohort has been described elsewhere (10, 11).

Our datasets originate from the PROTECT’s Product Use Survey, which study participants fill out at three separate follow up pregnancy visits, occurring at 16-, 20- and 24-weeks’ gestation (+/- two weeks). The survey consists of a series of questions regarding the use of 14 different personal care and household products designed to quantify possible exposure to chemical substances. In addition to the yes or no questions that assess if a participant used a certain consumer product. Those responding yes to a specific personal care product are asked to write all the specific brand names used as an open-ended question and stored as a text variable.

### Data transformation

We develop a process to transform open-ended questions into coded categorical variables. The first step focused on fixing structural errors of the brand names, which included naming conventions, misspellings, incorrect capitalization, and Spanish nomenclature of brand names that often lead to mislabeled categories (e.g., “Brand” could also be spelled “brand” or “BRAND”). To address these inconsistencies, we used Pandas *‘unique()’* function to visualize the unique and different naming conventions of the brand names. Since these values have a string data type, we used string methods to match and rename them to a single consistent brand name that resulted in a single categorical variable that contained a unique code for all reported brand names for each specific consumer product studied.

The second step addressed was the structural data characteristic that some participants had multiple entries of brand names for each product they used, per visit. Therefore, we needed to separate these multiple entries using the string *‘split()’* function that splits around a passed delimiter, of which our brand names included comma, “and”, “y” (Spanish for “and”), and ampersand (&). The returned data frame with all split strings was used to create an array of columns (e.g. SOAP97_1, SOAP97_2, SOAP97_3, etc.) and the original variable (SOAP97) is dropped using the *‘drop()’* function. Afterwards, we created a variable to count how many brands each participant report using at each of three scheduled visits, and we transformed this new data frame into a one-hot encoding data frame using Pandas *‘get_dummies()’* function that converts the categorical variables into indicator variables (dichotomous variables that represent if the participant used that particular brand). The result is a cleaner data frame that is primed for analysis.

Once we cleaned and structured the data set, we conducted a preliminary analysis of the data to understand the distribution of most commonly used brands (Figure 2). We encountered a high number of brand names and a highly skewed brand use distribution where a few brands had high use and accounted for most of the brand use. Therefore, we developed a selection process to identify what we called commonly used brands, defined as brands that retained their use rank throughout the three time periods of data collection. We used four criteria to select commonly used brands: 1) high total sample size (total number of participants that used all the brands selected), 2) high sample size per brand (number of participants that used each brand), 3) high number of brands per product and 4) brand usage remaining consistent throughout the three visits. This commonly used brand selection helped mitigate the effect that selecting all brands for the analysis would have on our sample size. By selecting a brand with low sample size, any future test would be statistically inconclusive (Kang et. al 2008).

### Method evaluation

To evaluate commonly used brand selection, we used Cohen’s power calculations for two-sample t-tests as described in the ‘pwr.t2n.test’ function from the ‘pwr’ package for R (version 1.3.0). Considering a moderate effect size (Cohen’s d) of 0.5, each test will include the highest ranked brand and the lowest ranked brand among the commonly used brands per product and visit. The highest ranked brand has the highest use frequency in contrast with the lowest ranked brands, which has the lowest use frequency from the selected brands. To better visualize the relationship of the highest ranked brand and the lowest ranked brand, a ratio was calculated to measure how much larger is the sample size from the highest ranked brand. We also compared how the commonly used brand selection differ per product in terms of the number of brands it selected, the total sample size and the power estimate by calculating a correlation matrix and analyzing the relationship between power, commonly used brands and brand usage.

## Results

The Product Use Survey collected data for 19 different personal care products, of which only 14 products had enough data to conduct the commonly used brand selection (Table 1). The products without sufficient data were excluded from this analysis. The product with the most brands was perfume with 331 different brands and the product with the least number of brands was shaving cream with 44 brands. The overall average for all studied products was 115 brands per product. The range of commonly used brands selected was between 2 (fabric softener and hair spray) and 8 (deodorant).

**Table1.** Commonly used brands frequency by visit.

| Type | Product | Total |  |  | Visit 1 |  | Visit 2 |  | Visit 3 |  |
| --- | --- | --- | --- | --- | --- | --- | --- | --- | --- | --- |
|  |  | Commonly used brands | Not commonly used brands | Used brands | Commonly used | Not commonly used | Commonly used | Not commonly used | Commonly used | Not commonly used |
| Personal care | Bar soap | 6 | 93 | 99 | 1216 (89.9%) | 136 (10.1%) | 1182 (89.9%) | 133 (10.1%) | 1032 (90.1%) | 113 (9.9%) |
|  | Conditioner | 4 | 149 | 153 | 603 (69.3%) | 267 (30.7%) | 549 (66.6%) | 275 (33.4%) | 521 (71.3%) | 210 (28.7%) |
|  | Deodorant | 8 | 71 | 79 | 1363 (91.9%) | 120 (8.1%) | 1497 (91.7%) | 136 (8.3%) | 1178 (92.5%) | 96 (7.5%) |
|  | Hair spray | 2 | 74 | 76 | 243 (58.4%) | 173 (41.6%) | 234 (59.8%) | 157 (40.2%) | 220 (63%) | 129 (37%) |
|  | Liquid soap | 6 | 98 | 104 | 1041 (88.4%) | 137 (11.6%) | 1101 (88.1%) | 149 (11.9%) | 928 (89.4%) | 110 (10.6%) |
|  | Lotion | 3 | 209 | 212 | 441 (53.3%) | 387 (46.7%) | 465 (54.3%) | 392 (45.7%) | 397 (53.8%) | 341 (46.2%) |
|  | Makeup | 6 | 102 | 108 | 343 (71.0%) | 140 (29%) | 306 (68.2%) | 143 (31.8%) | 290 (69%) | 130 (31%) |
|  | Nail polish | 3 | 46 | 49 | 195 (81.6%) | 44 (18.4%) | 197<br>(82.8%) | 41 (17.2%) | 158<br>(83.2%) | 32 (16.8%) |
|  | Perfume | 3 | 331 | 334 | 360 (35.7%) | 647 (64.3%) | 391<br>(38.7%) | 620<br>(61.3%) | 314<br>(35.8%) | 563 (64.2%) |
|  | Shampoo | 6 | 146 | 152 | 689 (78.3%) | 191 (21.7%) | 638 (76%) | 201 (24%) | 586<br>(80.8%) | 139 (19.2%) |
|  | Shaving cream | 3 | 41 | 44 | 60 (63.2%) | 35 (36.8%) | 62 (57.9%) | 45 (42.1%) | 54 (60%) | 36 (40%) |
| Home cleaning | Cleaning detergent | 6 | 68 | 74 | 677 (83.2%) | 137 (16.8%) | 635<br>(81.5%) | 144<br>(18.5%) | 620<br>(84.8%) | 111 (15.2%) |
|  | Fabric softener | 2 | 44 | 46 | 571 (77.2%) | 169 (22.8%) | 534 (78%) | 151 (22%) | 485<br>(76.7%) | 147 (23.3%) |
|  | General use cleaner | 6 | 74 | 80 | 406 (91.2%) | 39 (8.8%) | 439<br>(90.1%) | 48 (9.9%) | 398<br>(89.6%) | 46 (10.4%) |

As seen in Table 1, the most used product was deodorant, averaging 1463 participants per visit, while the least used product was shaving cream with an average of 97 participants that used it per visit. Coincidently the product with the highest commonly used brands, deodorant, reported the highest percentage of selected brand use per visit at around 92%. In contrast, one of the products with the lowest commonly used brands, fabric softener, reported a high percentage of selected brand use per visit at around 77.3% when compared to other products with more commonly used brands such as makeup; fabric softener reported around 69.4% of selected brand use with only six selected brands.

Table 2 shows the ratio and power when evaluating commonly and not commonly used brands by visit. From all products, fabric softener, conditioner, and lotion where the products that attained the highest power with the lowest ratio. Makeup was the following product with a high power in the first visit, but a score of 0.49 and 0.54 was attained for the second and third visit.

**Table 2.** Power analysis of two sample t-test comparing the highest ranked commonly used brand and the lowest ranked commonly used brand by visit.

| Type | Product | Visit 1 |  |  |  | Visit 2 |  |  |  | Visit 3 |  |  |  |
| --- | --- | --- | --- | --- | --- | --- | --- | --- | --- | --- | --- | --- | --- |
|  |  | Max | Min | Ratio | Power | Max | Min | Ratio | Power | Max | Min | Ratio | Power |
| Personal care | Bar soap | 523 | 32 | 16.3 | 0.78 | 505 | 45 | 11.2 | 0.89 | 420 | 38 | 11.1 | 0.83 |
|  | Conditioner | 268 | 73 | 3.7 | 0.97 | 254 | 54 | 4.7 | 0.91 | 223 | 62 | 3.6 | 0.93 |
|  | Deodorant | 412 | 46 | 9 | 0.89 | 417 | 40 | 10.4 | 0.85 | 348 | 28 | 12.4 | 0.72 |
|  | Hair spray | 202 | 41 | 4.9 | 0.83 | 193 | 41 | 4.7 | 0.83 | 172 | 48 | 3.6 | 0.86 |
|  | Liquid soap | 479 | 21 | 22.8 | 0.61 | 498 | 20 | 24.9 | 0.59 | 424 | 12 | 35.3 | 0.4 |
|  | Lotion | 220 | 60 | 3.7 | 0.93 | 260 | 47 | 5.5 | 0.88 | 221 | 50 | 4.4 | 0.89 |
|  | Makeup | 79 | 26 | 3 | 0.59 | 79 | 19 | 4.2 | 0.49 | 72 | 23 | 3.1 | 0.54 |
|  | Nail polish | 153 | 14 | 10.9 | 0.43 | 168 | 12 | 14 | 0.38 | 134 | 10 | 13.4 | 0.33 |
|  | Perfume | 226 | 53 | 4.3 | 0.90 | 254 | 56 | 4.5 | 0.92 | 197 | 50 | 3.9 | 0.88 |
|  | Shampoo | 264 | 41 | 6.4 | 0.84 | 250 | 34 | 7.4 | 0.78 | 221 | 37 | 6 | 0.8 |
|  | Shaving cream | 33 | 10 | 3.3 | 0.27 | 34 | 13 | 2.6 | 0.32 | 25 | 12 | 2.1 | 0.28 |
| Home<br>cleaning | Cleaning<br>detergent | 342 | 29 | 11.8 | 0.73 | 294 | 36 | 8.2 | 0.81 | 293 | 37 | 7.9 | 0.82 |
|  | Fabric softener | 438 | 133 | 3.3 | 0.99 | 408 | 126 | 3.2 | 0.99 | 354 | 131 | 2.7 | 0.99 |
|  | General use<br>cleaner | 171 | 4 | 42.8 | 0.17 | 156 | 4 | 39 | 0.17 | 152 | 6 | 25.3 | 0.22 |

Max, Sample size of highest ranked commonly used brand; Min, Sample size of lowest ranked commonly used brand; Ratio, Ration between Max and Min.

In 1S Table, the correlation coefficient between the commonly used brand frequency of each visit approximated 0.99. Similarly, there was a moderate positive relationship between unique commonly used brands and ratio with a coefficient of 0.44 for visit 1, 0.42 for visit 2 and 0.47 for visit 3. However, we observed a moderate inverse relationship between the total unique brands and the total brand usage. Finally, we found a moderate inverse relationship (r < -0.56) between the ratio and the power for all three visits.

## Discussion

The differences in brand use distributions per product provided an optimal environment for evaluating the performance of the commonly used brand selection methodology. The variance of power calculated is not attributed to number of commonly used brands selected, but to frequency of use of the selected brands themselves. We observed that products with similar total unique brand and total unique commonly used brands, produced very different power. Shaving cream and fabric softener had similar commonly used brands, total used brands and ratio, but vastly different commonly used brand frequency, thus leading to the gap in calculated power as presented in Table 2. Bar soap and liquid soap also had similar commonly used brands but different commonly used brand frequency, yet their calculated power only deviated 0.03.

The commonly used selection methodology selected a relatively low amount of commonly used brand from some products like fabric softener (n = 2) and hair spray (n = 2). This can be attributed to the difference in brand use distribution per product. However, this difference in commonly used brands between products doesn’t translates to low sample size. The percentage of participants that used the commonly used brands for fabric softener was 77.2% in the first visit. Similar percentage was observed in makeup with 71% for the first visit and for that product the number of commonly used brand was six. The power for makeup and fabric softener was 0.59 and 0.99 respectively as described in Table 2. This suggests that even if the methodology selects only a small number of brands, it makes up for it in performance of future statistical tests.

The lowest performer regarding power was general use cleaner with a power of less than 22 through the three visits (Figure 1S). This product has similar characteristics of use as cleaning detergent, expect for the ratio between the highest ranked brand and the lowest ranked brand amongst the commonly used brands. During post-collection analysis, selecting low sample size exposure groups provides difficulty in establishing significant research outcomes, therefore, using the commonly used brand selection methodology as is for this product might not yield noteworthy results. However, products like perfume who performed well with a power of around 0.90 and six commonly used brands selected, in terms of the commonly used brand frequency for the three visits it was 36.7%; suggesting that the number of commonly used brand selected could be increased without affecting the legitimacy of the outcomes.

**Figure 1.**
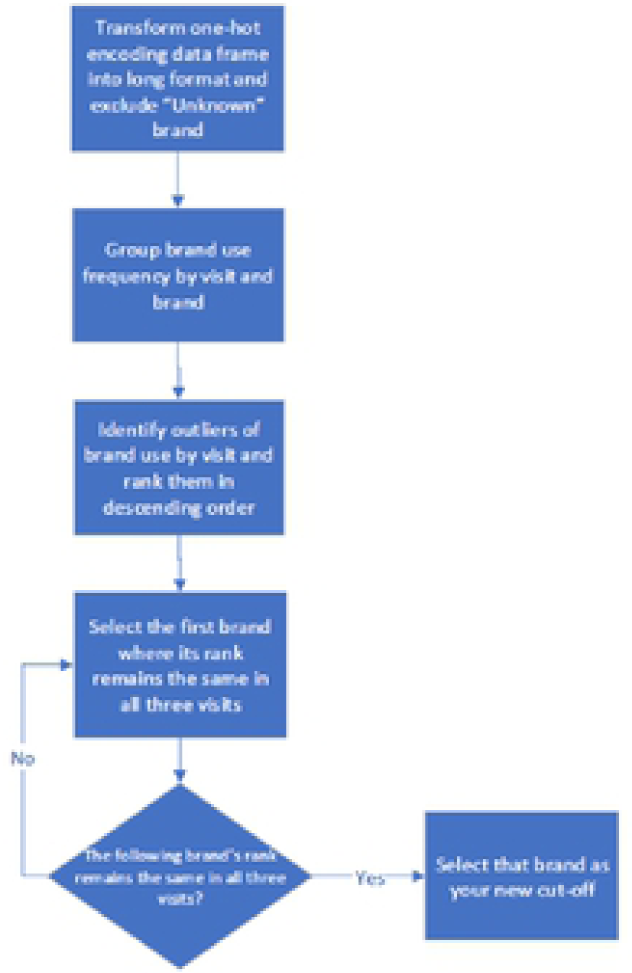
Commonly used brand selection flow chart.

This methodology can be easily applied to any data set that records brand use in repeated measures and contribute to the development of analysis plans that involve open-ended questions in product use questionnaires and any other exposure related forms. However, this methodology cannot be applied to cross-sectional studies (one data point per participant) since the algorithm requires multiple measures to establish the framework of what is a commonly used brand. This limitation is compensated by the fact that not having multiple measures, sampling issues are not present, therefore allowing to select brands based on optimal sample size.

A limitation of this process is that with open-ended questions some responses include the company’s name rather than the brand used. This may cause inaccuracies regarding the measurement of EDCs exposure for that specific brand since companies often sell a type of personal care product under multiple brands. This phenomenon, as explained by Berger (15), could lead to a skewed reporting towards companies with high number of brands.

The commonly used brand selection methodology allows researchers to objectively analyze their data and not select brands for analysis based on subjective or arbitrary conditions. While ranked selection methods exist in other fields, the Borda count (16) in political science for example, we found no literature on brand selection methodologies for environmental exposure analysis. Further studies using different distributions are needed to test the reproducibility of this methodology within public health and environmental research.

## Data Availability

All relevant data are within the manuscript and its Supporting Information files.

## Acknowledgements

We are extremely grateful to all PROTECT study participants and their families. The authors also thank the nurses and research staff who participated in cohort recruitment and follow up, as well as the Federally Qualified Health Centers (FQHC) in Puerto Rico that facilitated participant recruitment, including Morovis Community Health Center, Prymed in Ciales, Camuy Health Services, Inc. and the Delta OBGyn Group in Manatí, as well as the Manatí Medical Center and the Metro Pavía Hospital in Arecibo.

## Funding

This research is supported by grants from the National Institute of Environmental Health Sciences (P42ES017198) and the Environmental Influences on Child Health Outcomes (5UH3OD023251 and 3UH3OD023251-05S1) ECHO is a nationwide research program supported by the NIH, Office of the Director to enhance child health.

The research protocol was approved by the Ethics and Research Committees of the University of Puerto Rico, the University of Michigan School of Public Health, University of Georgia, and Northeastern University. All study activities were fully explained to each participant before obtaining written consent for participation in the study.

